# SARS-CoV-2 infections in kindergartens and associated households at the start of the second wave in Berlin, Germany – a cross sectional study

**DOI:** 10.1101/2020.12.08.20245910

**Authors:** Marlene Thielecke, Stefanie Theuring, Welmoed van Loon, Franziska Hommes, Marcus A. Mall, Alexander Rosen, Falko Böhringer, Christof von Kalle, Valerie Kirchberger, Tobias Kurth, Joachim Seybold, Frank P. Mockenhaupt, on behalf of the BECOSS study group

## Abstract

**Objectives:** The comparatively large proportion of asymptomatic SARS-CoV-2 infections in the youngest children opens up the possibility that kindergartens represent reservoirs of infection. However, actual surveys in kindergartens beyond individual outbreaks are rare. At the beginning of the second pandemic wave in Berlin, Germany, i.e., end of September 2020, we screened SARS-CoV-2 infections among kindergarten children, staff and connected household members.

**Methods:** Twelve kindergartens were randomly selected in the Berlin metropolitan area, and a total of 720 participants were recruited (155 pre-school children, 78 staff, 487 household members). Participants were briefly examined and interviewed, and SARS-CoV-2 infections and anti-SARS-Cov-2 IgG antibodies were assessed.

**Results:** Signs and symptoms, largely resembling common cold, were present in 24.2% of children and 28.9% of staff. However, no SARS-CoV-2 infection was detected among 701 PCR-tested individuals, and only one childcare worker showed IgG seroreactivity (0.15%; 1/672).

**Conclusions:** Against a backdrop of increased pandemic activity in the community, this cross-sectional study does not suggest that kindergartens are silent transmission reservoirs. Nevertheless, at increasing pandemic activity, reinforced precautionary measures and repeated routine testing appears advisable.

## Introduction

Measures to contain the SARS-CoV-2 pandemic particularly affect kindergartens and schools, many of which were closed in the early phase of the pandemic. The debate as to whether kindergartens are safe for children and educators continues [1,2]. However, actual data on infection prevalence and transmission in such facilities are scarce [3-5]. Here, we aimed at assessing the prevalence of SARS-CoV-2 infections and IgG seroreactivity among pre-school children, educators, and household members connected with Berlin kindergartens during the beginning of the second pandemic wave.

## Methods

Twelve Berlin kindergartens were visited in this cross-sectional study between September 28 and October 2, 2020. In that week, 1,561 PCR-confirmed SARS-CoV-2 infections were registered in Berlin (29 and 43 cases in children aged 0-4 and 5-9 years of age, respectively) [6], the 7-day-incidence was 38 cases/100.000 inhabitants [7], and numbers started to grow exponentially (Figure 1). For the selection of twelve of >2700 kindergartens in Berlin, the city districts were divided into three strata according to socio-economic status. In each stratum, two districts were randomly selected using a random number generator, and in each selected district, two kindergartens were randomly chosen. Nine kindergartens unwilling to participate were replaced by resampled substitutes following the above-mentioned selection procedure. At each kindergarten, we aimed at recruiting 20 children and 5 staff, whenever possible, belonging to one care-group. Household members of children and staff were invited to also participate in the survey. Informed written consent was obtained from all participants (by legal representatives in the case of minors), and the study was reviewed by the ethics committee of Charité – Universitätsmedizin Berlin (EA2/091/20). All procedures respected the Declaration of Helsinki, 2013 revision.

**Figure 1.**
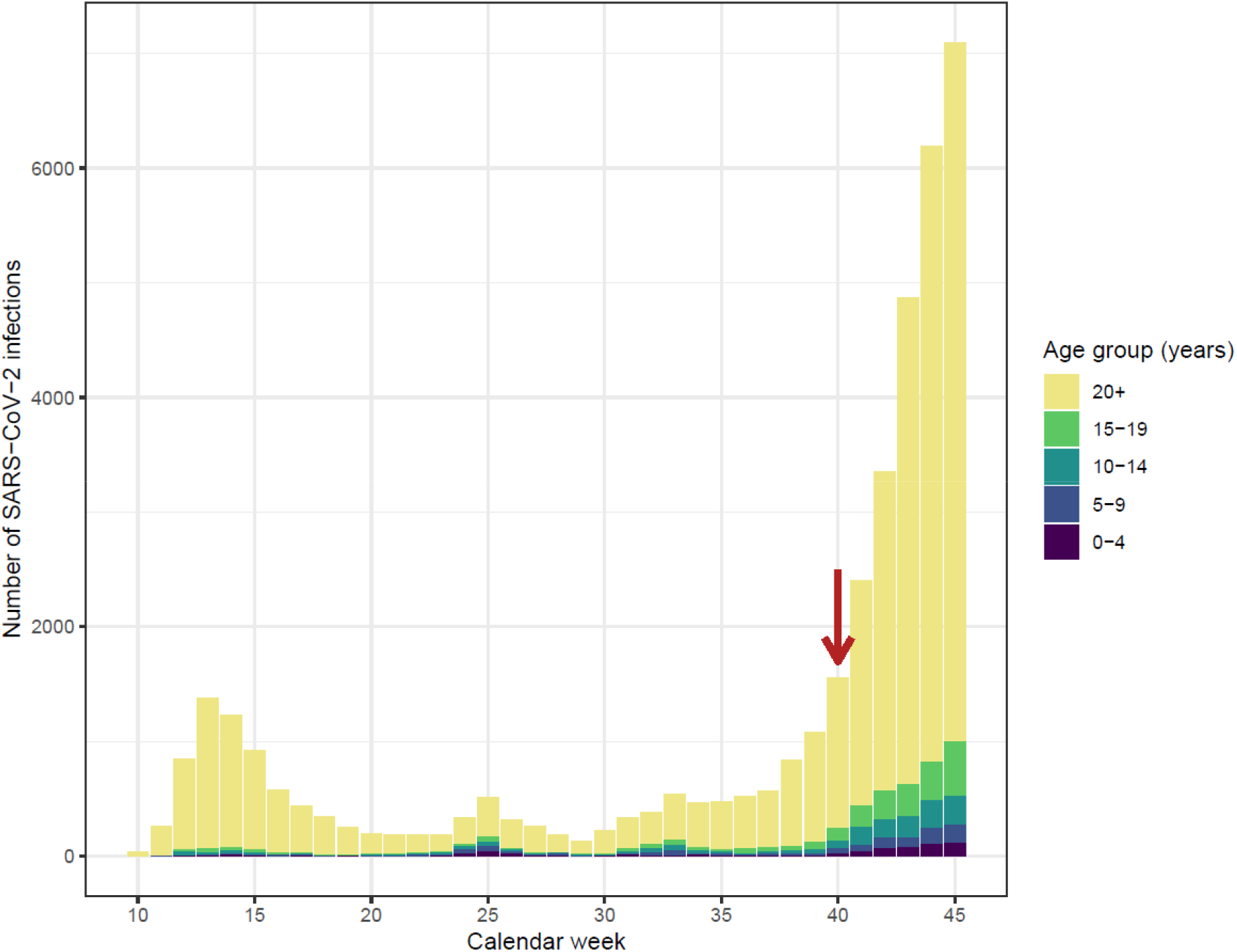
Notified, PCR-confirmed SARS-CoV-2 infections in Berlin *per* calendar week Note. The study period is indicated by a red arrow. Data are derived from [9].

A study team visited each facility to collect samples and data. Children and staff were interviewed on present signs and symptoms, and forehead temperature was measured. Swabs (eSwab, Copan, Italy) were taken from the oropharynx and nasal vestibules [8]. Household members were invited during pick-up time to mobile clinics set up off the kindergartens. They brought their self-collected swabs (oropharynx and nasal vestibules) after having received an illustrated instruction and swabs beforehand. Finger-prick blood samples were collected on filter paper (BioSample Card, Ahlstrom Munksjö, Germany). Five reportedly ill participants were visited at home. SARS-CoV-2 infection was determined by real-time-PCR using the cobas^®^ SARS-CoV-2 test on cobas^®^ 6800/8800 systems (Roche Diagnostics, Switzerland). For the assessment of anti-SARS-CoV-2 IgG, 4.75 mm discs were punched from dried blot spots, samples were extracted in 250 µl sample buffer at ambient temperature for one hour, and ELISA was performed on a EUROLabWorkstation (Euroimmun AG, Germany).

Participants and household members completed a digital questionnaire covering signs and symptoms in the preceding two weeks, risk factors, and hygiene behaviour-related measures. Moreover, infection prevention measures implemented at the kindergartens were documented.

## Results

In total, 720 individuals participated in the study (155 pre-school children, 78 staff, 487 household members). The median age of the kindergarten children was 4.4 years; 40% were girls. Educators were predominantly mid-aged females (Table 1). Two in three household members were adults (68.6%, 334/487; median age, 39 years; range, 21 – 90) in addition to schoolchildren (20.3%; 99/487) and younger children (10.5%, 51/487). Signs and symptoms were present in one in four kindergarten children, including runny nose (17.0%, 26/153), cough (11.1%, 17/153), and sore throat (2.0%, 3/153). Leading complaints among symptomatic educators (28.9%) were headache (14.7%, 11/75), runny nose (13.5%, 10/74), and cough (11.8%, 9/76). Upon examination, 2.6% and 1.3% of children and educators were febrile, respectively (Table 1). Any chronic condition was stated for 6.2% (8/130) of children, 38.2% (26/68) of educators, and 17.6% (66/374) of household members. Most kindergarten children reportedly never wore a facemask, with substantially higher use rates among educators and household members (Table 1).

**Table 1.** Characteristics of kindergarten children, educators, and household members

| Characteristic | Children | Educators | Household members |
| --- | --- | --- | --- |
| No. (%) | 155 (21.5) | 78 (10.8) | 487 (67.6) |
| Age, years (median, range) | 4.4 (1.0 – 6.3) | 44 (18 – 78) | 36 (0 - 90) |
| Female, % (n/n) | 40.0 (60/150) | 86.8 (66/76) | 47.8 (229/479) |
| Any present symptom, % (n/n) | 24.2 (37/153) | 28.9 (22/76) | n.a. |
| Present fever ($\geq 37.5^{\circ}\text{C}$), % (n/n) | 2.6 (4/152) | 1.3 (1/76) | n.a. |
| Any symptom in preceding two weeks, % (n/n) | 57.0 (73/128) | 74.6 (50/67) | 58.4 (215/368) |
| Never wears facemask, % (n/n) | 97.8 (90/92) | 34.4 (22/64) | 23.0 (76/330) |

n.a., not available

As for regulations by the Berlin senate implemented in the kindergartens, two-thirds (8/12) of the facilities had a physical distance rule between staff and 91.7% (11/12) between staff and parents. Obligatory facemask wearing by staff was stated in 41.7% (5/12) for parental contacts and in 8.3% (1/12) for contacts among colleagues. Attendance despite (afebrile) symptoms of common cold was allowed by 75% (9/12) of daycare centres, and 72.7% (8/11) of kindergartens reported daily training of hygiene rules with the children. In most facilities (83.3%, 10/12), children were taken care of in fixed groups and rooms, which were ventilated more than five times/day in half of the cases.

Swabs could be collected for 98.1% (152/155) of children, all educators (78), and 96.7% (471/487) of household members. In none of these 701 samples, SARS-CoV-2 was detected. Only one childcare worker showed IgG seroreactivity (0.15%; 1/672), with an optical density value of 1.5 (threshold, >1.1).

## Discussion

As compared to adults, children <10 years of age appear to be less frequently infected with SARS-CoV-2, to show rather mild disease or asymptomatic infection, and may be less infectious [2,3,9,10]. However, actual data from kindergarten settings are scarce. During 3 months of low incidence, no SARS-CoV-2 infection was seen among 825 kindergarten children with weekly buccal mucosa swabs in Hesse, Germany [4]. While our survey took place during increased pandemic activity in Berlin, reflected by 2 to >10 times higher 7-days-incidence than in the before-mentioned study [11], we still observed the absence of SARS-CoV-2 infection in kindergarten children, staff, and household members. Although reassuring, these findings do not exclude the possibility that community infections spill over into daycare settings.

Data from Australia and elsewhere indicate that infections in daycare centres mirror the degree of community transmission [12]. During the first pandemic wave in New South Wales, Australia, ten kindergartens were affected by SARS-CoV-2 infections, with one case *per* facility. No secondary transmission occurred except for one of the ten facilities where an adult primary case gave rise to an outbreak. The secondary attack rate in educational settings, including schools, was 1.2% (excluding the single outbreak, 0.4%) but lower in child-to-child contacts (0.3%) than in staff-to-staff contacts (4.4%) [3], endorsing presumed low child-to-child transmission [3,5]. Almost inevitably, outbreaks in daycare facilities occur [3,5], including in one of our studied kindergartens at the end of November 2020. In this regard, asymptomatic, thus, undetected infections may pose a risk of initiating infection chains in kindergartens even though viral loads are substantially lower in asymptomatic than in symptomatic pre-school children [13]. A large-scale seroprevalence study among Bavarian children showed that the majority of cases is missed, that children below and above seven years exhibit similar proportions of seroreactivity, and that nearly half of the infections had been asymptomatic [14]. Against this background, standard precautionary measures need to be re-enforced at increasing pandemic activity to reduce transmission in kindergartens. Rapid response to detected cases, including subsequent contact tracing and quarantine, is essential. However, considering the frequent asymptomatic course of SARS-CoV-2 infection in young children, this may be insufficient. Recent modelling data suggest that repeated screening, e.g., twice per week, combined with instant reporting, isolation, or quarantine, can greatly reduce viral transmission in a population. In contrast, test sensitivity is of minor importance [15]. This suggests implementing regular screening, e.g., by antigen test, in kindergarten settings, possibly self-administered to disburden the currently overstretched healthcare sector. Notwithstanding, upcoming repeat examinations in the study kindergartens will help elucidating the role of daycare centres in transmission at varying incidences.

## Data Availability

Data are available with the authors.

## Transparency declaration

The authors declare that they have no conflict of interest.

This study was supported by the Senate of Berlin. The funder had no role in study design, data collection and analysis, decision to publish, or preparation of the manuscript.

## Authors’ contributions

MT and ST organized data collection, data management, and analysis. WvL did statistical analysis, data collection. FH set up and maintained the database set-up. MAM and AR supervised paediatric examinations. FB did the laboratory examinations. CvK supervised database and logistics. VK supervised data collection and liaised with health and educational authorities. TK did sample selection and data analysis. JS organized staff allocation and logistic issues. FPM designed and implemented the study, contributed to data collection and analysis. BECOSS study group did data collection. All authors participated in drafting the article or revising it critically for intellectual content.

## BECOSS study group members

Tanja Chylla, Elisabeth Linzbach, Annkathrin von der Haar, Jennifer Körner, Maximilian Gertler, Julian Bernhard, (Institute of Tropical Medicine and International Health; Charité – Universitätsmedizin Berlin); Heike Rössig, Marco Kurzmann, Frederike Peters, Christoph Wiesmann, Johanna Horn, Julia Steger, Norma Bethke (Medical Directorate, Charité – Universitätsmedizin Berlin); Tobias Schmidergall, Frederik Holz, Antje van den Berg, Maria Luz Peña-Groth (Department of Pediatric Pulmonology, Immunology and Critical Care Medicine, Charité – Universitätsmedizin Berlin)

We thank the kindergartens and the respective staff for enabling the study in their facilities.

